# Survey on use of Colistin in poultry and its residues in Farm- fresh and commercial eggs in Sabon-gari Kaduna State, Nigeria

**DOI:** 10.1101/2025.11.20.25340666

**Authors:** Lauratu Lawal Halilu, Junaidu Kabir, Grace Kia, Oche Anthony Ameh, Saaaondo James Ashar

## Abstract

Antimicrobial microbial resistance (AMR) is a priority global public health threat. It is driven by increased used of antimicrobial agents in humans, veterinary medicine and agriculture resulting in selection pressure and occurrence of their residues in edible products. It is essential to preserve critical antibiotics such as colistin which is a last resort drug for treatment of MDR Gram-negative bacterial infection. A total of 1,180 eggs were collected over a period of 3 months from both farms and markets in Sabon-gari local government area of Kaduna State Nigeria to determine the prevalence of colistin and other antibiotic residues. The eggs were pooled in batches of 10 and screened for antimicrobial residues using microbial inhibition test, positive samples were further subjected to a confirmatory test using Elabscience® colistin ELISA kit. Structured questionnaire was administered to farmers to obtain information of antimicrobial usage, intensity of colistin uses, purpose of use and other management information, awareness on drug withdrawal period, antibiotic residue and their effects and implication on public health. Out of the 118 pooled samples, 75.4% were positive for antimicrobial residue out of which 37.5% were due to colistin residue. Farms that reared less than 500 birds had higher levels (50%) of colistin residue in their eggs as compared to those that reared between 500-4999 (37.5%). Farmers who used antibiotics weekly and those who did not observe withdrawal period had higher levels (56.5%) of colistin in their eggs. There was association between presence of colistin residue and non-observance of withdrawal period. Keproceryl^®^ was the most frequently used antibiotic by farmers in this study. This study further confirms the presence of antimicrobial residues particularly colistin in both farm-fresh and commercial eggs within Sabon-gari local government area and the need for regulations to monitor drug administration by farmers.

## 1.0 INTRODUCTION

Poultry are reared all over the world for meat and egg production. Egg production constitutes a large proportion of poultry output. Egg is a major food source, providing good quality balanced nutrients to billions of people throughout the world. The world’s total egg production exceeded 86.3 million metric tons in 2021 from 73.9 million metric tons in 2016 (Shahbandeh, 2023). Poultry production in Nigeria amounts up to 300 million tonnes of meat and 650 million eggs annually (FAO, 2019). The current level of production cannot meet the demand posed by a rapidly growing population, thereby demanding for intensification of production. Drugs as an essential part of poultry production are used to prevent and control diseases, reduce stress due to environmental changes, vaccination and other management practices (Kabir *et al.,* 2004). Veterinary drug use is accompanied by concern for the persistence of the drugs as residues. Drug residues present in foods are known to constitute global health concerns. Such health problems include the development of allergic reactions, emergence of multiple resistant strains of pathogenic bacteria (Clewell, 1981; Ogarawa, 1981; Gomes and Demoly, 2005), interference with spermatogenesis, development of cancer and mutation in humans (Ali *et al*., 1987; NAFDAC, 1996; Dollery, 1999; FAO, 2003).

Colistin, also termed as polymyxin E, is a hydrophilic polycationic peptide which was discovered in 1949 and then introduced for the treatment of gram-negative bacteria such as *Acinetobacter* species, *Pseudomonas aeruginosa*, *Klebsiella* species and *Enterobacter* species (Komura and Kurahashi, 1979; Falagas and Kasiakou, 2005). The direct mechanism of action of colistin is to cause damage of the outer membrane, which triggers the leakage of cell contents and subsequent cell death (Newton, 1956; Schindler and Osborn, 1979).

Recently, colistin is being used as last-line treatment against Gram-negative bacterial infections (Bergen *et al*., 2015). However, the development of multiple-drug resistance in numerous bacteria, especially Gram-negative bacteria has limited the treatment options for various bacterial infections resulting in the re-emergence of cyclic polypeptide drugs like colistin remains the only option.

The effectiveness of currently available antibiotics is decreasing due to increasing number of resistant bacterial strains causing infections (Ventola, 2015). Over the years, the issue of antibiotic residues in edible animal products and their effect on human health has been a significant concern (Bahry *et al*., 2013).

Food animals have been determined to be the reservoir for colistin resistance microbes from which they spread to other hosts (Skov and Monnet, 2016). Due to the adverse effects of colistin, many developed countries have prohibited its usage in food animals, but developing countries are still in use of colistin in food animal production, thereby imposing a major public health risk. As a result, there is need for surveillance to detect and quantify the use of antimicrobials in poultry and the occurrence of colistin residues in order to implement effective measures towards control.

## 2.0 MATERIALS AND METHODS

### 2.1 Sampling of Eggs

The study was a cross sectional study involving two target populations: farm-fresh eggs from farms and commercial eggs marketed in the area for the detection of colistin. The sampling points were selected based on inclusion and exclusion criteria.

### 2.2 Questionnaire Survey

A total of 59 structured questionnaires were administered to the layer poultry farms and to broiler and other producers in Sabon Gari Local Government Area of Kaduna State order to obtain information on antimicrobial usage, intensity of use of colistin, purpose for use and other management information. Also, the questionnaire assessed the awareness of the farmers on drug withdrawal period, antibiotic residues and their effects and the implications on public health.

### 2.3 Testing of Egg for Antimicrobial residue

The antimicrobial screening of eggs was carried out using the disc diffusion method where *Bacillus subtilis* ATCC 6633 was used as test organism. An 18-hour culture of the test organism in10mls nutrient broth (Oxoid Basingstoke, Hampshire, U.K) was used to inoculate the plates. The plates were inoculated with test organism by dipping sterile cotton swab sticks into the suspension of the test organism until it was saturated with the suspension. The Mueller Hinton agar plates were then uniformly swabbed, one quadrant at a time as described by Granite Diagnostics (1985), to achieve a lawn of confluent growth. Egg yolk was tested by dipping a sterile 1.2cm diameter Whatmann filter paper disc into the test yolk sample until the disc was saturated with yolk sample. The impregnated disc was gently placed on the surface of the Mueller Hinton agar plate inoculated with the test organism. The disc was gently and firmly pressed on the surface to allow for proper diffusion of the agar around the disc. The plates were then incubated at 37^0^C for 18-20 hours. After incubation, the plates were viewed for the presence or absence of zones of inhibition to test organism around test and control discs. The diameter of the zones of inhibition was measured in millimetres from one edge of the zone to the other across the disc. The difference between the diameter of zone of inhibition and that of the disc was calculated. Any disc with a difference of 1mm or more was considered positive.

The positive samples were further tested using a commercial test kit, Colistin–specific competitive enzyme immunosorbent assay (c-ELISA) -Elabscience for the quantitative analysis of colistin in the matrices was used to detect colistin residue in eggs. The optical density (OD Value) for each well was determined at 450 nm with a microplate reader which was done 5 minutes after stop reaction.

## 3.0 RESULTS

A total of 92(78%) pooled egg samples out of the 118 pooled egg samples collected tested positive for antimicrobial residues using microbial inhibition test. When further tested with a Colistin- ELISA kit only 34 (39.5%) egg samples tested positive for colistin residues. Farms that did not observe antibiotic withdrawal period had a higher prevalence 13 (28.3%), there was a statistically significance association between occurrence of colistin residue in egg and failure to observance of withdrawal period (Chi square Value ꭓ^2^ = 4.394, P = 0.036, OR= 0.271).

**Table 1:** Prevalence of Colistin Residues in Eggs from Markets and Farms Using Microbial Inhibition Test (MIT) in Sabon Gari Local Government Area of Kaduna State.

| Location | Number Examined | Number Positives (%) |
| --- | --- | --- |
| **Markets** |  |  |
| Samaru | 29 | 24 (20.3) |
| Sabon-gari | 30 | 18 (15.3) |
| **Farms** |  |  |
| Samaru | 42 | 37 (31.4) |
| Sabon-gari | 17 | 13 (11.0) |
| **Total** | **118** | **92 (77.97)** |

**Table 2:** Prevalence of Colistin Residues in Eggs from Markets and Farms Using Colistin ELISA in Sabon Gari Local Government Area of Kaduna State.

| Location | Number Examined | Number Positive (%) |
| --- | --- | --- |
| Markets | 40 | 17 (19.8) |
| Farms | 46 | 17 (19.8) |
| **Total** | **86** | **34 (39.5)** |

Out of the 59 questionnaire that were administered to the famers, majority were between the age of 20-39 (45.1%), 26 (44.1%) were between 40-59 while (3.4%) respondents fell between the age 60 and above. The educational status of the farmers showed that majority (78%) had tertiary education and 1 respondent (1.7%) had informal education. The respondent (33.9%) had 3-6 years of poultry experience,3.4% respondent had < 3years farm experience. Majority of the existent farms (44.1%) were between 3-6 years, 33.9% had > 10 years of experience and (15.3%) had < 3year experience. The study showed that there were no significant differences (p>0.05) between the frequency of drug administration and the experience of poultry farming.

Thirty- three (33) out of the 59 farmers interviewed kept both layers and broilers in their farms (59.9%) while 26(44.1%) kept layers. Twenty-eight 28(47.5%) had a flock size of <500 birds and birds between 500-4999. Deep litter system was the most predominate management system (74.6%). Majority of the farms (71.2%) used borehole water as a source of drinking water. The type of feed used was mostly commercial feed (83.1%). Twenty-nine (49.2%) of farmers used antibiotics rarely used antibiotics on demand followed by those that used once a month 19 (32.2%) and those that used weekly 10 (16.9%). The treatment of bacterial infection was the major reason (49.2%) of antibiotic usage which was followed by use for prevention of infection (23.7%) while (5.1%) used antibiotics for growth and egg production. Most farmers (54.2%) observed drug withdrawal period, while others (45.8%) failed to observe drug withdrawal period. Drug recommendation on most of the farms (72.9%) was done by a veterinary doctor while drugs recommended by self were (18.6%). Some farmers (25.4%) sometimes used gloves while administering antibiotics while (37.3%) farmers never used gloves, with majority of farmers (84.7%) always washing their hands after antibiotic administration.

**Table 4.3:**
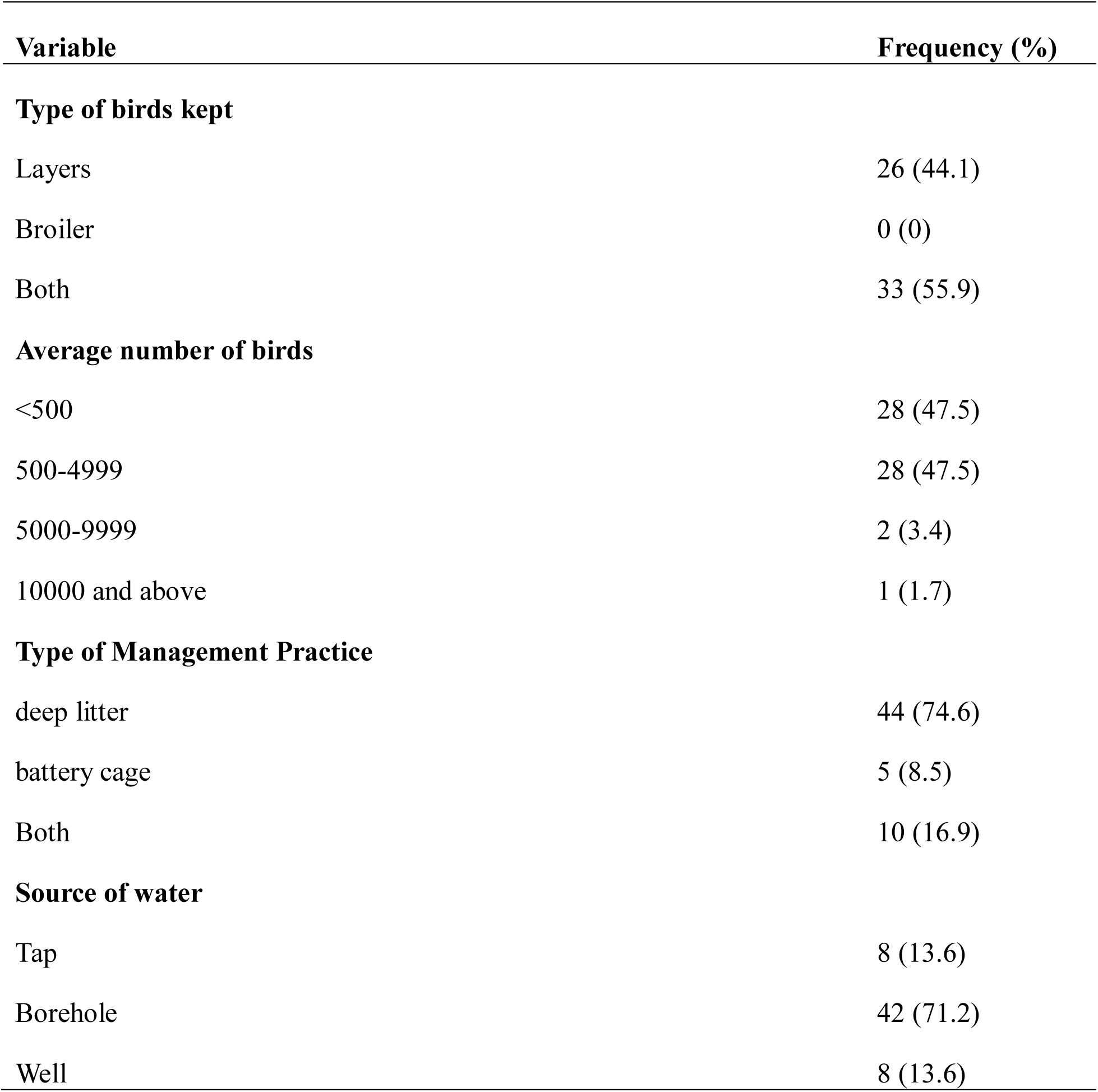
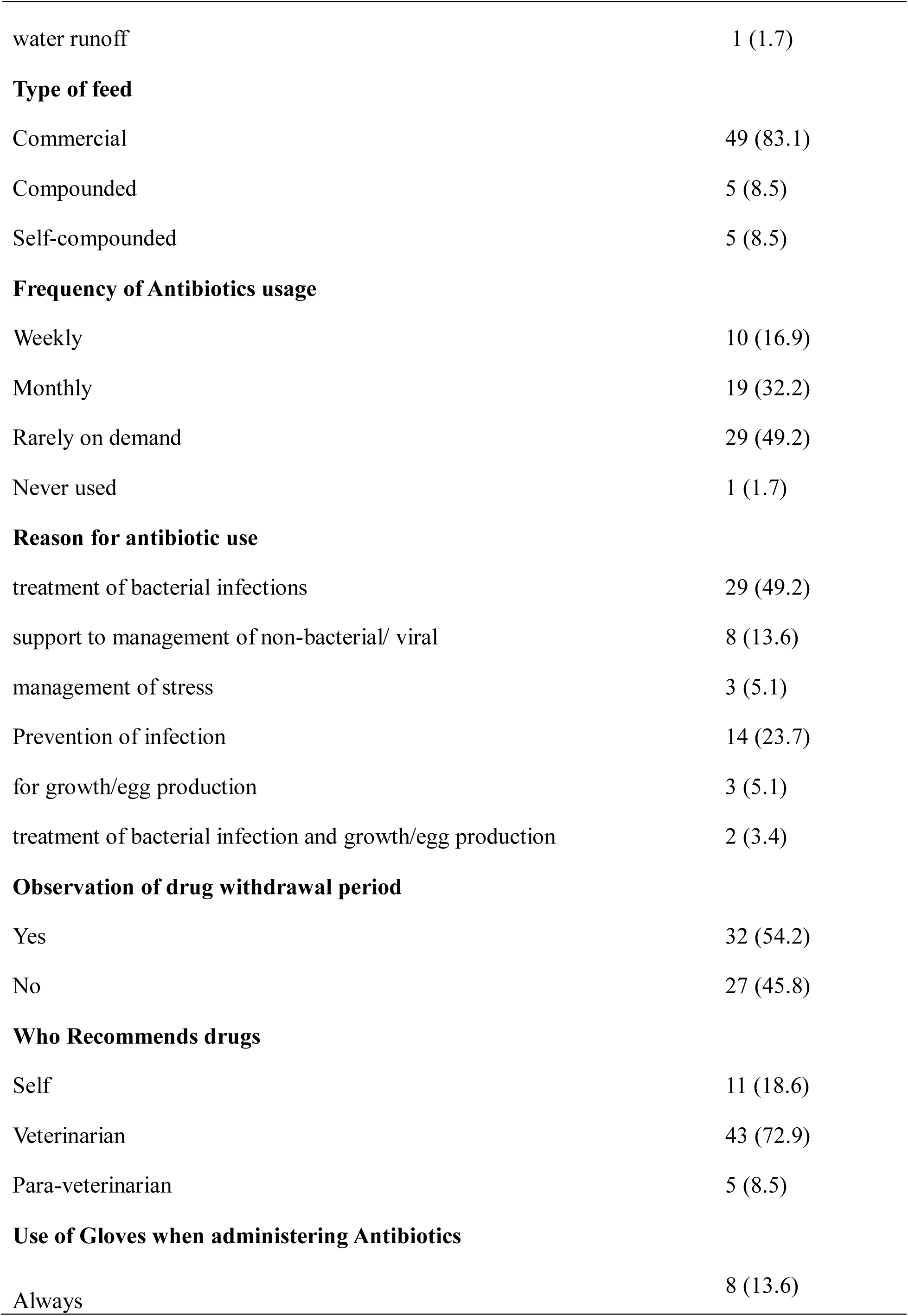
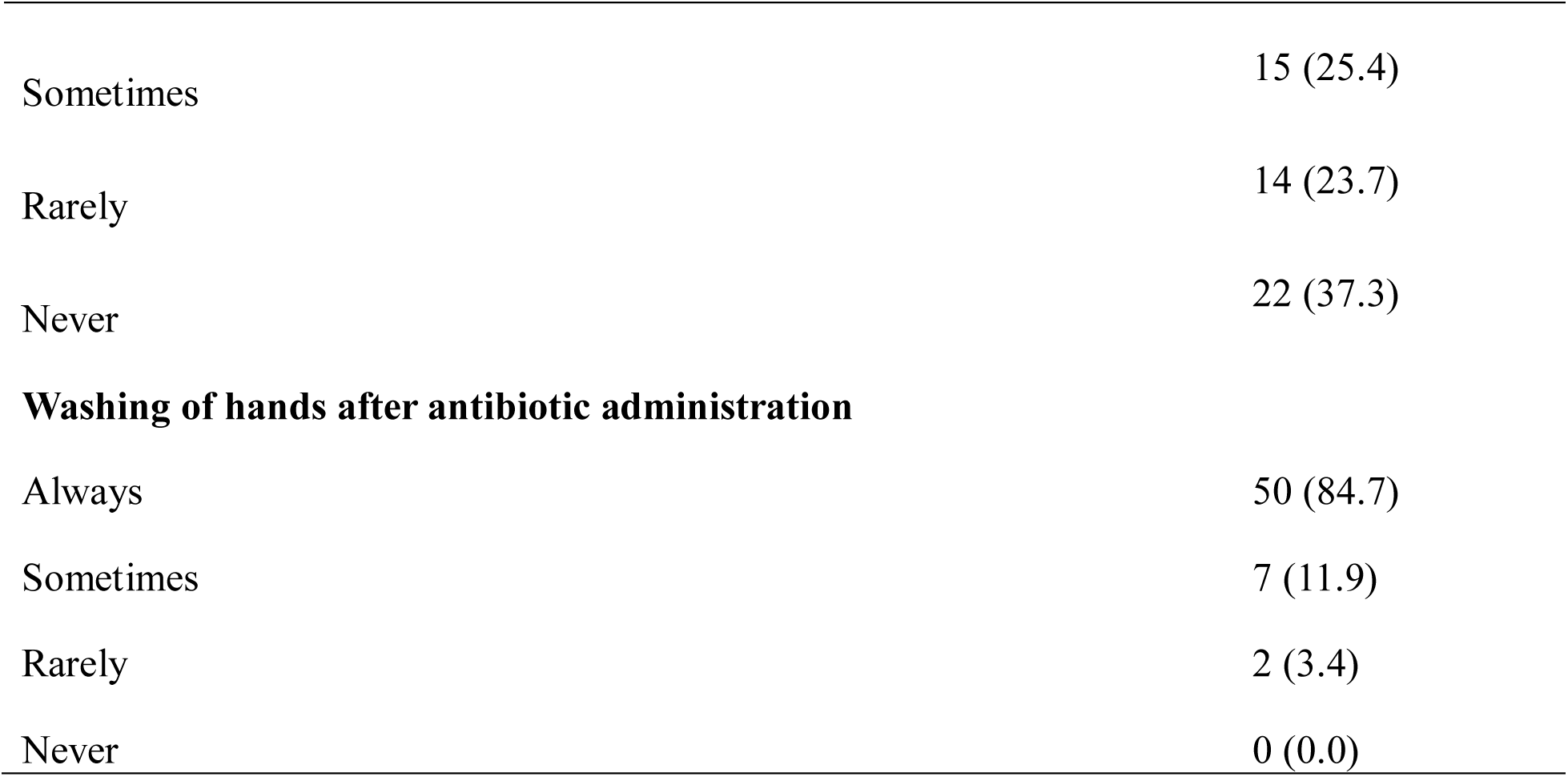
Practices of Poultry Farmers with Respect to Antibiotic Use in Farms in Sabon Gari Local Government Area of Kaduna State, Nigeria.

| Variable | Frequency (%) |
| --- | --- |
| **Type of birds kept** |  |
| Layers | 26 (44.1) |
| Broiler | 0 (0) |
| Both | 33 (55.9) |
| **Average number of birds** |  |
| <500 | 28 (47.5) |
| 500-4999 | 28 (47.5) |
| 5000-9999 | 2 (3.4) |
| 10000 and above | 1 (1.7) |
| **Type of Management Practice** |  |
| deep litter | 44 (74.6) |
| battery cage | 5 (8.5) |
| Both | 10 (16.9) |
| **Source of water** |  |
| Tap | 8 (13.6) |
| Borehole | 42 (71.2) |
| Well | 8 (13.6) |
| water runoff | 1 (1.7) |
| **Type of feed** |  |
| Commercial | 49 (83.1) |
| Compounded | 5 (8.5) |
| Self-compounded | 5 (8.5) |
| **Frequency of Antibiotics usage** |  |
| Weekly | 10 (16.9) |
| Monthly | 19 (32.2) |
| Rarely on demand | 29 (49.2) |
| Never used | 1 (1.7) |
| **Reason for antibiotic use** |  |
| treatment of bacterial infections | 29 (49.2) |
| support to management of non-bacterial/ viral | 8 (13.6) |
| management of stress | 3 (5.1) |
| Prevention of infection | 14 (23.7) |
| for growth/egg production | 3 (5.1) |
| treatment of bacterial infection and growth/egg production | 2 (3.4) |
| **Observation of drug withdrawal period** |  |
| Yes | 32 (54.2) |
| No | 27 (45.8) |
| **Who Recommends drugs** |  |
| Self | 11 (18.6) |
| Veterinarian | 43 (72.9) |
| Para-veterinarian | 5 (8.5) |
| **Use of Gloves when administering Antibiotics** |  |
| Always | 8 (13.6) |
| Sometimes | 15 (25.4) |
| Rarely | 14 (23.7) |
| Never | 22 (37.3) |
| **Washing of hands after antibiotic administration** |  |
| Always | 50 (84.7) |
| Sometimes | 7 (11.9) |
| Rarely | 2 (3.4) |
| Never | 0 (0.0) |

### Knowledge of Poultry Farmers with Regards to Antibiotics

Majority of the respondents were well informed and knowledgeable about antibiotics. Majority of the respondents (86.4%) knew about antibiotic withdrawal period and knew that it should be observed (81.4%). On the other hand, some respondents (45.8%) did not know that eggs obtained from treated birds could harbour residue. Most respondent (62.7%) did not agree that eggs laid by farm birds during antibiotic administration be sold out. A good number of respondents (69.5%) are aware tha antibiotics boost egg laying. A good number of respondents (67.8%) were not aware of the public health effect of antibiotic residue. Also, majority of the respondents (84.7%) did not know of the Government regulation on antibiotic usage. Most respondents (62.8%) also knew antibiotic use results in drug residue.

### Attitude of Poultry Farmers to Antibiotic Usage

The attitude of respondents with respect to antibiotic use (Table 4.10) showed that majority (18.90±4.35) agreed and felt that antibiotic could be used as treatment drug. Most (14.20± 3.77) of the respondents felt antibiotic could be used as a prophylactic drug, while others (11.47±3.39) felt antibiotic should be used as a growth promoter. Very few (5.10±2.26) farmers believed that eggs laid by sick birds should be consumed immediately. Some farmers (9.68± 3.11) believed antibiotic usage in birds results in deposits of residues in eggs while other farmers (11.42±3.38) felt antibiotic residue is of public health significance. Other farmers (10.25± 3.20) agreed that residues result following antibiotic usage. Most farmers (13.86±3.72) believed that the practice of withdrawal period would leave no antibiotic residues in eggs. A few farmers (6.19±2.49) were of the opinion that antibiotic residue in egg is beneficial to human health.

### Frequently Used Antibiotic on Farms

A total of 57 farms out of the 59 farms sampled, used antibiotics. Most of the antibiotics used by the farmers are sold as cocktail of antibacterial agents. Ten antibacterial classes were used on the farms (Table 4.13) with Colistin (59.6%), Oxytetracycline (50.6%), Doxycycline (42.1%) and Streptomycin (40.4%) being the highest used in combination. The least used antibiotic agents include Sulfonamides and Chlorampenicols in combinations respectively. Metronidazole 0(0%) was the least used antibiotic in combined formulation while Enofloxacin 3 (5.3%) was the most used antibiotic as a single formulation.

**Table 4:** Frequently Used Antibiotic on Farms in Sabon Gari Local Government Area of Kaduna State, Nigeria.

| Class of Antibiotics | Active ingredients | Number of farms with single ingredients (%) | Number of farms with combined ingredients (%) | Number of farms not using the active ingredient | Total number of farms using antibiotics |
| --- | --- | --- | --- | --- | --- |
| Beta-lactames | Amoxicillin | 0(0) | 9 (15.8) | 48 | 57 |
| Tetracyclines | Oxytetracycline | 1 (1.8) | 30 (50.6) | 26 | 57 |
|  | Doxycycline | 0 (0) | 24 (42.1) | 33 | 57 |
| Aminoglycoside | Gentamicin | 0 (0) | 19 (33.3) | 38 | 57 |
|  | Neomycin | 0 (0) | 2 (3.5) | 55 | 57 |
|  | Streptomycin | 1 (1.8) | 23 (40.4) | 33 | 57 |
| Macrolides | Erythromycin | 0 (0) | 18 (31.6) | 39 | 57 |
|  | Tylosin | 0 (0) | 9 (15.8) | 48 | 57 |
| Chloramphenicol<br>s | Chloramphenicol | 0 (0) | 2 (3.5) | 55 | 57 |
|  | Florphenicol | 1 (1.8) | 3 (5.3) | 53 | 57 |
| Sulfonamides | Sulphadimidine | 0 (0) | 2 (3.5) | 55 | 57 |
|  | Sulpharidine | 0 (0) | 3 (5.3) | 54 | 57 |
|  | Sulphaquinoxaline | 1 (1.8) | 2 (3.5) | 54 | 57 |
|  | Sulphamerazine | 0 (0) | 3 (5.3) | 54 | 57 |
|  | Trimethoprim | 0 (0) | 2 (3.5) | 55 | 57 |
|  | Sulphadiazine | 0 (0) | 4 (7.0) | 54 | 57 |
| Quinolones | Enrofloxacin | 3 (5.3) | 12 (21.1) | 42 | 57 |
|  | Ciprofloxacin | 0 (0) | 7 (12.3) | 50 | 57 |
| Polypeptides | Colistin | 0 (0) | 34 (59.6) | 23 | 57 |
| Nitroimidazole | Metronidazole | 1 (1.8) | 0 (0) | 56 | 57 |
| Nitrofurans | Furaltadone | 0 (0) | 8 (14.0) | 49 | 57 |
| Amprolium | Amprolium | 0 (0) | 2 (2.5) | 55 | 57 |

### Antibiotics in use in Sabon-gari Local government and their trade names

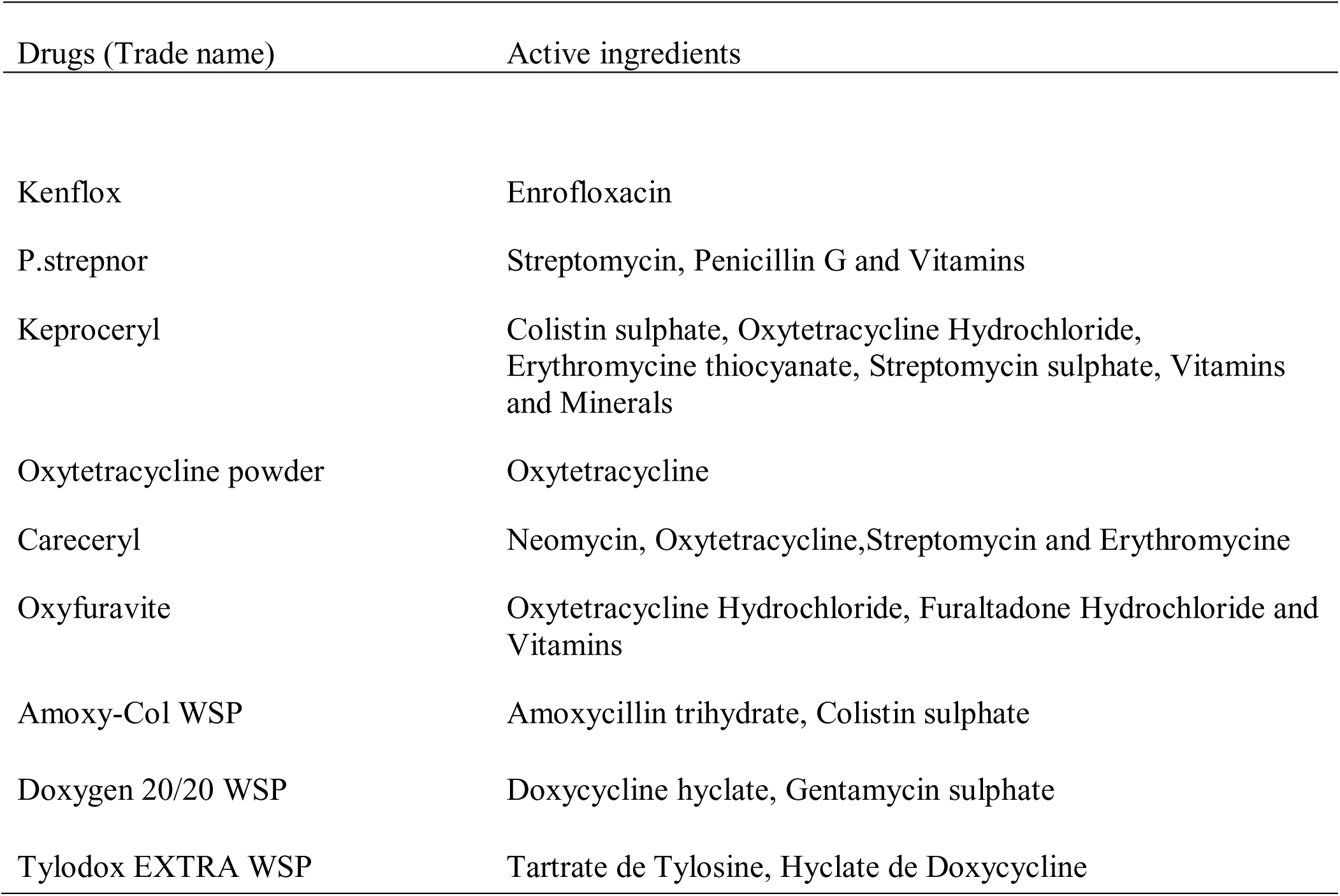
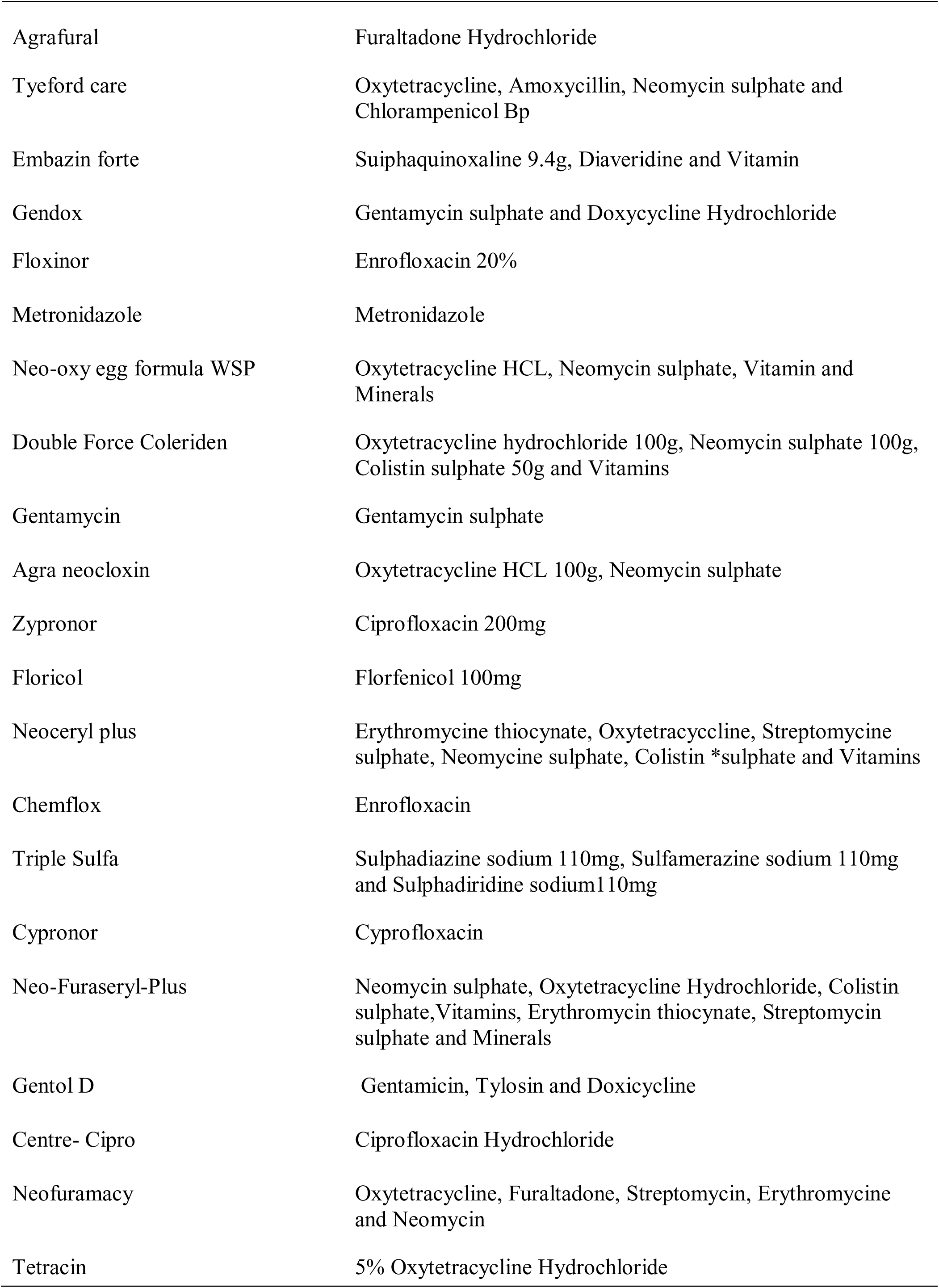
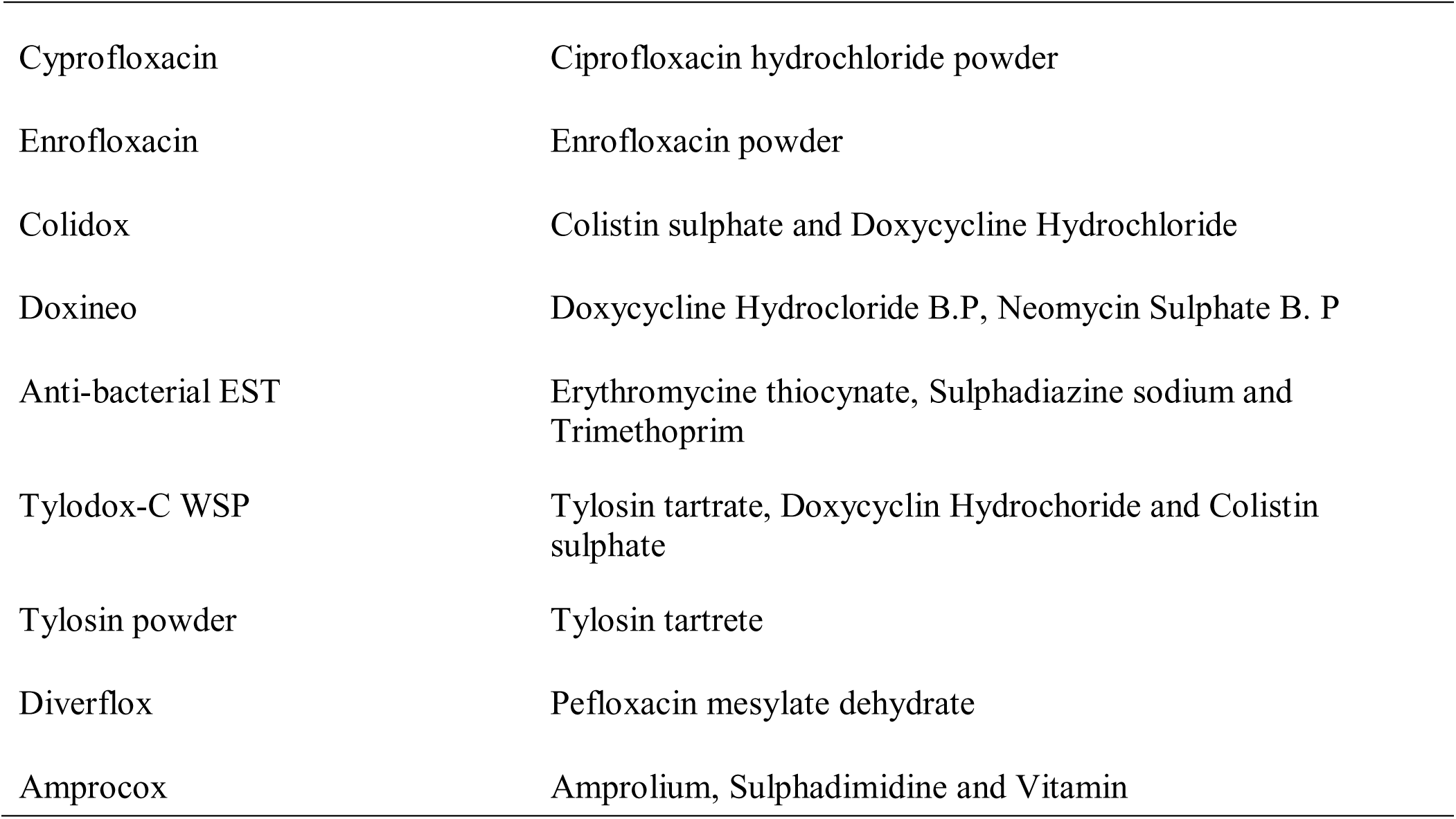

## Discussion

This study investigated the prevalence of antibiotic residues as well as colistin residues in poultry products in the Zaria. Egg samples in this study were obtained from a rapidly growing poultry area located in the major agricultural region of northern Nigeria. Veterinary drugs are easily obtained over the counter in markets and veterinary shops. The prevalence of antimicrobial drug residues obtained in this survey (78%) indicates a high incidence of veterinary drug residues in eggs of chicken in Sabon gari local government Area, Zaria. The occurrence of antimicrobial drug residues found in the study is considerably high when compared with previous studies carried out by Fagbamila *et al*. (2010) and Omeiza *et al*. (2012) who reported the occurrence of 3.6% and 7.6% respectively. This perhaps signifies a commensurate high level of use of antimicrobials, since all the farmers studied indicated that they use antibiotics.

Colistin is used in poultry production to treat important Gram-negative bacteria such as *Eschericia coli* and *Salmonella* and in most cases, is frequently abused (Qin *et al.,*2017). This may be related to the high prevalence (39.5%) of its residues in this survey. This may indicate that colistin is considered as the last-line treatment of multidrug-resistant gram-negative bacterial infections (Qin *et al.,*2017). The occurrence of colistin residues in eggs was previously reported by Sirdar *et al*. (2012) and Oluwasile *et al*. (2014) who reported the use of oxytetracycline, colistin, tylosin and enrofloxacin in poultry farms. Despite the restrictions (EMA, EU and WHO) associated with the use of this antibiotic, it is excessively being used by farmers to treat enteric diseases and for growth promotion (Kumar *et al.,* 2020).

Poultry farming, particularly broiler and layer, is considered the hotspot in the use of antimicrobials for therapeutic and non- therapeutic purposes. It is thought that antimicrobial misuse, abuse and overuse are common practices in poultry rearing across the world (Laxminarayan *et al.,* 2016). Consequently, the overuse and misuse of colistin led to the development of colistin resistance as reported in previous studies (Wongsuvan *et al.,* 2018). It was also observed that farmers ignorantly used the same antibiotic but under different brand names, with colistin residues found in eggs intended for human consumption. Results from this study showed that 39.5% of the egg samples tested positive for Colistin residues using competitive ELISA. The detection limit for the kit is 20ppb with one sample having 299.419ppb, close to the maximum residual limit for colistin (CX/MRL 2-2021). The egg sample with the highest residues level was obtained from a small-scale farm (≤500 birds which lacked basic biosecurity measures hence a major public health concern.

The study area is characterized by high demand for poultry products, especially eggs, which has attracted many households towards backyard poultry production with possible consequences of uncontrolled usage of drugs (Omeiza *et al.,* 2012). The high prevalence of colistin residue (21.96%) observed in this study for small scale/backyard poultry farms <500 was relatively higher than large scale farms which may be indicative of improved biosecurity in the large-scale farms. This strongly indicates that small scale farmers are more prone to frequent drug use due to greater likelihood of exposure of the birds to infections.

Results of this study also showed that 72.6% of the farms frequently sought professional consultation from a veterinarian in the management of poultry diseases, out of which 31.0% used these drugs without following veterinary recommendations. The consequence of not heeding to professional recommendations often results in abuse or misuse of antibiotics with resultant accumulation of residues in animal products.

The frequency of colistin usage was highest in farms that administer antibiotics monthly (15.2%). These findings are similar to those previously reported by Adebowale *et al*. (2008) and Geidam *et al*. (2012). This is confirmed by the opinion of some respondents (45.8%) who have knowledge that eggs obtained from treated birds could harbour drug residues. There was no association between frequency of usage of colistin and the occurrence of colistin residues in this study. This may be due to the fact that even single use could result in residues, particularly where withdrawal period is not observed.

Even though the findings in this study showed that nearly half of the farmers did not observe the recommended withdrawal period (10 days) while some participants were unaware of the term “withdrawal period”. There was an association between failure to observe of withdrawal period and occurrence of colistin residues in the study. This practice of not observing withdrawal period by farmers increases the possibility of high level of antibiotic residues with detrimental health consequences to the public. These findings are similar to reports by Al-Mustapha *et al*. (2020) and Guetiya *et al*. (2016) who stated that non-adherence to withdrawal period is of public health concern as antimicrobial residue in food producing animals may lead to multiple drug resistance pathogens in humans. This lack of adherence to withdrawal periods is largely because farmers are avoiding losses and sell off birds when it is economically favourable to them. This is enabled by lack of enforcement by regulatory bodies. Many researchers, including Annan-Prah *et al*. (2012) and Mubito *et al*. (2014), reported that the problem of residues is partly driven by economic factors in addition to lack of awareness. Farmers fear losses associated with and discard of contaminated poultry products due to residues where legislations are weak. This work supports the findings of other researchers, who reported antibiotic residues in poultry and poultry products in Nigeria (Idowu *et al*., 2010 and Kabir *et al.,* 2004).

This study evaluated the knowledge, attitude, and practices (KAP) of poultry farmers regarding antibiotic use, antibiotic residues, most commonly used antibiotics, reason for use of antibiotics, drug withdrawal period, reason for use of antibiotics, frequency of antibiotics use, type of feed, source of water and types of management practiced. The study described demographic information of farmers, including farm existence and years of experience. Most 27 (45.8%) poultry producers in our study were aged below 40 years old and the highest percentage 46 (78%) of farmers were university graduates presumably they should be well informed about the public health concerns like residues. Most 26 (44.1%) of the poultry farmers had an average of 3-6 years experience in poultry farming, therefore should have a good understanding of legislations regarding poultry production in Nigeria. Overall, the demographic picture of the poultry farmers was similar with some former studies by Geidam *et al*. (2018), Al-Mustapha *et al*. (2020).

The study revealed that most 43 (72.9%) poultry farmers used antibiotics prescribed by veterinarians, which differs from the study of Omeiza *et al*. (2012), Okonkwo *et al*. (2019) and Fagbamila *et al*. (2010) where more than 33.3% of the farmers used antibiotics without the prescriptions of the veterinarians. This could be attributed in part to the fact that pharmaceutical representatives sell drugs directly to farmers. Sometimes, to promote the sale of antibiotics, pharmaceutical companies provide their own veterinary assistance through dealers to small farmers. There are results of promotion of animal drug usage by pharmaceutical companies and their agents to farmers in other countries such as China, Nepal, India etc (Roess *et al*., 2013). Most 29 (49.2%) farmers used antimicrobials for therapeutic and prophylactic purposes in this study. WHO/WOAH categorized antibiotics into three (3) classes which includes veterinary highest priority critically important antimicrobials agents (which colistin belongs), veterinary highly important antimicrobial agent and veterinary important antimicrobial agents. This study showed that farmers believed that prophylactic use of antibiotics could save veterinary costs and treatment costs by reducing the prevalence of diseases (Okeke *et al*., 2005). However, this practice does not conform with the Muscat declaration of 2023 on the use of antibiotic which targets at reducing total antimicrobial use in agrifood systems, ensuring that ‘access’ group antibiotics represent at least 60 per cent of overall antibiotic consumption in humans by 2030 as well as preserving critically important antimicrobials for human medicine and ending the use of important antimicrobials for growth promotion in food animals (Global conference on AMR, 2022).

Furthermore, farmers in this study believed that preventive usage of antimicrobials might reduce the disease burden and stop frequently occurring poultry diseases especially where vaccination against poultry diseases has not been carried out. This is similar to reports by Sirdar *et al*. (2012) and Roess *et al*. (2013). However, this is in contrast to laid down regulations by WOAH (2021) which categorizes colistin as one of the highest priority reserve group of antibiotics which should not be used as a prophylactic or first treatment unless justified. Therefore, extra-label use should be limited and its use as growth promoter should be urgently prohibited (WOAH, 2021). Also, the prophylactic use of antimicrobials as recorded in this study may result from lack of knowledge and wrong advice from the drug vendors.

In this study, only 3 (5.1%) of farmers reported that they use antibiotics for growth promotion. Farmers did not report the use of antimicrobials as growth promoters in the study of Nure Alam *et al*. (2022) which is similar to this study where only 2 (3.4%) used antibiotics for growth and egg production. This varies with the findings of Olatoye (2011) who reported that 86% of poultry farms in Ibadan, Nigeria used antibiotics for growth promotion. Preventive use of antibiotics in commercial poultry production is also common in other Asian countries, including Cambodia, Indonesia, Vietnam and Thailand (Coyne *et al*., 2019).

This study assessed the varying perception of the farmers towards antibiotic use and occurrence of residues.The various use of antibiotics by poultry farms in this survey have also been reported by other authors such as Ogunleye *et al*. (2008) who reported the high usage of enrofloxacin, tetracycline, gentamicin streptomycin, furatadone, tylosin, norfloxacin among poultry farms in Abeokuta, while Sirdar *et al*. (2012) also reported the use of oxytetracycline, colistin, tylosin and enrofloxacin among poultry farms in Khartoun, Sudan, Oluwasile *et al*. (2014) and Nure Alam *et al*. (2022) reported high usage of enrofloxacin, colistin, neomycin and gentamicin. These findings are in agreement with the observations in Zaria by this study where colistin, erythromycin, tetracycline, doxycycline, gentamicin, streptomycin and enrofloxacin are the commonly used. Among these, ciprofloxacin and tylosin and colitstin are considered ‘Highest Priority Critically Important Antimicrobials’ for public health (WHO, 2019).

As observed in this study, the widespread use of antibiotics can lead to the transfer of such drugs into the human food chain in form of drug residues in animal derived foods. This can occur when withdrawal periods are not observed before selling the products to the public for human consumption. It also leads to lower efficacy of such drugs when used for treatment of human illness. Besides, antimicrobial usage results to antibiotic resistant bacterial strains with the possibility of transfer of resistant genes to other pathogenic and non-pathogenic bacteria. Humans can thus acquire antimicrobial resistant bacteria following consumption of poultry products. It is for these reason that the WHO, FAO and WOAH consider poultry as the most important first point of surveillance for priority zoonotic bacterial pathogens resistant to antimicrobials. Antibiotic residues in food such as eggs as shown by this study, poses an indirect health risk. The World Health Organization (WHO) estimates antibiotic-resistant diseases to cause 10 million deaths each year by 2050. This will eventually destroy the economy leading to a similar situation like the 2008-2009 global financial crisis (Arsene *et al.,* 2021), therefore the occurrence of residues in eggs in Zaria may constitute only a small fraction of the problem in Nigeria.

The study also investigated the attitude of farmers to antimicrobial drug use and residues. Majority of the farmers agreed to strongly agreed that antibiotics should be administered frequently (9.88±3.14), should be used as growth promoters (11.47±3.339) or as prophylaxis (14.20±3.77). These indicates a bad perception amongst the farmers on the use of antibiotics. Most farmers disagree that; eggs laid by sick birds under medication should be consumed immediately (5.1±2.26), antibiotics usage in birds results in deposits of residues in eggs (9.68±3.11). Farmers also agreed that residues result following antibiotic usage (10.25±3.20), the practice of withdrawal period would leave no antibiotic residues in eggs (13.86±3.72). some farmers were uncertain that antibiotic residue is of public health significance (11.42±3.38), most farmers strongly disagreed that antibiotic residue in eggs is beneficial to human health (6.19±2.49) these indicates the good perception of the farmers toward antibiotic usage. Good attitude amongst of the farmers will be favourable for public health awareness and control of drug residues in poultry products.

## Conclusion

The study showed a high prevalence of antimicrobial drug residues in eggs from farms and markets using microbial inhibition test was 92(78%), as well as a high prevalence of colistin residues 34(39.5%) in poultry eggs. Colistin is widely used with its residue occurring in high concentration in an egg sample from a farm (up to 299.419ppb). Consumers in the study area may be constantly exposed to violative levels of these residues. There was a statistically significant association (Chi square Value ꭓ^2^ = 4.394, P = 0.036, OR= 0.271) between farms that fail to observe withdrawal period and occurrence of residues. The most frequently used antibiotic was Colistin (59.6%) with farms using the drug as Keproceryl® at the time of the study which suggests that the alarming high prevalence antibiotics residues (colistin) results from indiscriminate use of the antibiotics by farmers in the study area. There is need for a strong awareness on the dangers of these practices should be created by the relevant bodies while government should strengthen the existing controls on the production, distribution and use of these drugs especially the banned ones. There is also the need to have a national program to monitor the presence of antimicrobial residues and their metabolites in poultry and other animal products meant for human consumption. Policies should be on ground which enforce and encourage small scale poultry farms to be registered by Government to enhance better practice of biosecurity in order to reduce drug abuse and misuse. The contribution of antimicrobial drug residues in edible animal tissue to emergence and spread of resistant pathogens in Nigeria should be further investigated.

## Data Availability

All data in the present study are available upon reasonable request to the authours

